# Decoding natural gait cycle in Parkinson’s disease from cortico-subthalamic field potentials

**DOI:** 10.1101/2022.05.02.22274438

**Authors:** Kenneth H. Louie, Ro’ee Gilron, Maria S. Yaroshinsky, Melanie A. Morrison, Julia Choi, Coralie de Hemptinne, Simon Little, Philip A. Starr, Doris D. Wang

**Affiliations:** Department of Neurological Surgery, University of California San Francisco, San Francisco, CA, 94143, USA; Department of Radiology, University of California San Francisco, San Francisco, CA, 94143, USA; Department of Applied Physiology & Kinesiology, University of Florida, Gainesville, FL 32611, USA; Department of Neurology, University of Florida, Gainesville, FL, 32608, USA; Department of Neurology, University of California San Francisco, San Francisco, CA, 94143, USA

**Author notes:** Correspondence to: Doris D. Wang, MD, PhD, 505 Parnassus Ave, M779, San Francisco, CA 94143-0112.

## Abstract

Human bipedal walking is a complex motor behavior that requires precisely timed alternating activity across multiple nodes of the supraspinal network. However, understanding the neural dynamics that underlie walking is limited. We investigated the cortical-subthalamic circuit dynamics of overground walking from three patients with Parkinson’s disease without major gait impairments. All patients were implanted with chronic bilateral deep brain stimulation leads in the subthalamic nucleus (STN) and electrocorticography paddles overlying the primary motor (M1) and sensory (S1) cortices. Local field potentials were wirelessly streamed through implanted bidirectional pulse generators during overground walking and synchronized to external gait kinematics sensors. We found that the STN displays increased low frequency (4-12 Hz) spectral power between ipsilateral heel strike to contralateral leg swing. Furthermore, the STN shows increased theta frequency (4-8 Hz) coherence with M1 through the initiation and early phase of contralateral leg swing. Our findings support the hypothesis that oscillations from the basal ganglia and cortex direct out-of-phase, between brain hemispheres in accordance with the gait cycle. In addition, we identified patient-specific, gait-related biomarkers in both STN and cortical areas at discrete frequency bands. These field potentials support classification of left and right gait events. These putative biomarkers of the gait cycle may eventually be used as control signals to drive adaptive DBS to further improve gait dysfunction in patients with Parkinson’s disease.

## Introduction

Human walking is a complex motor task that requires the flexible coordination of both cortical and subcortical structures within the brain. Natural upright walking consists of each leg alternating between the stance phase, when the foot is in contact with the ground, and the swing phase, when the foot is in the air; these two phases make up the “gait cycle,” comprised of a series of stereotyped, predictable events such as left and right heel strikes and toe offs. The left and right legs must maintain reciprocal, swing/stance phase coordination that is critical for stable bipedal gait.

The subthalamic nucleus (STN) and primary motor cortex are likely key nodes of the supraspinal network that regulates human gait, given the STN’s projection to the locomotor regions in the brainstem (Takakusaki, 2017), and its direct connections to the motor cortex via the hyperdirect pathway (Nambu et al., 2002). Understanding of the cortico-subthalamic network activities that underlie natural walking in humans is, however, limited due to methodological constraints. Studies using noninvasive methods such as scalp electroencephalography (EEG) studies have shown that natural healthy overground walking is associated with rhythmic increases and decreases in the alpha (8-12 Hz), beta (13-30 Hz), and gamma (70-90 Hz) frequency ranges from the sensorimotor cortical regions of healthy subjects (Gwin et al., 2011; Seeber et al., 2015; Wagner et al., 2012). Although, EEG lacks the spatial resolution to discern whether these rhythms originate from the motor cortex to coordinate locomotion or represent sensory feedback during walking, and are prone to movement artifacts. Basal ganglia field potentials recorded from implanted deep brain stimulation (DBS) leads of patients with Parkinson’s disease (PD) have also revealed modulation of beta (13-30Hz) oscillations from the subthalamic nucleus (STN) while stepping in place (Fischer et al., 2018; Hell et al., 2018; Tan et al., 2018) and during overground walking throughout the gait cycle (Arnulfo et al., 2018; Canessa et al., 2020; Hell et al., 2018). However, because aberrant beta oscillatory synchrony in the STN is a hallmark of akinesia in PD (Hammond et al., 2007; Little and Brown, 2014), and beta oscillations decreases with movement planning and execution in general, including those of the upper extremity (Eisinger et al., 2020; Kühn et al., 2004; Wingeier et al., 2006), whether these subthalamic beta modulations represent biomarker of specific gait events is unclear. Finally, little is known about cortical-subthalamic interactions during the natural gait cycle. Cortical EEG and STN LFP in PD patients with freezing of gait appear to show low frequency (4-13 Hz) synchrony during effective walking, and decouple during freezing episodes (Pozzi et al., 2019). However, the nature and role of these low frequency synchrony in effective walking is unknown.

Our hypothesis is that the STN interacts with the motor cortex in a temporal-specific manner to coordinate reciprocal leg movements to generate effective bipedal locomotion. We investigated the cortical-subthalamic circuit dynamics of natural walking from three patients with PD without major gait disturbances implanted with chronic bilateral STN DBS leads and motor cortex electrocorticography (ECoG) paddles. Neural oscillatory activities were simultaneously and wirelessly streamed from the bilateral primary motor (M1) and sensory (S1) cortices as well as the STN during overground walking, and these LFPs were synchronized to external gait kinematic sensors. Our aims were: 1) to characterize the oscillatory signatures of natural walking from the STN and sensorimotor cortices, 2) to identify cortico-subthalamic circuit coherence changes throughout the gait cycle, and 3) to determine accuracy of gait event decoding (i.e., heel strike or toe off) based on these cortical and subthalamic oscillatory signatures.

## Results

### STN shows coordinated low frequency power modulation during walking

To investigate STN and sensorimotor spectral changes during the gait cycle, we extracted gait cycle epochs from the continuous wavelet transform of a patient’s LFP data, divided the epochs into bins representing 1% of the gait cycle, and tested whether the power at a specific frequency during gait cycle differs from the average walking power at the same frequency. We found significant ventral and dorsal STN LFP spectral power modulations during the gait cycle, with the two hemispheres showing coordinated, reciprocal power changes. Grand average of all gait cycles across all subjects demonstrated increased amplitudes of alpha-gamma frequency (10-50 Hz) band power in the ventral STN and of low frequency (5-15 Hz) band power in the dorsal STN. These increases occurred during weight acceptance, the period from ipsilateral heel strike to contralateral toe off (Figure 1A and B, top). The left STN showed increased power during the double limb support period, between left heel strike to right initial swing (∼0-10% of the gait cycle), while the right STN showed this power increase during right heel strike to left initial swing (∼50-60% of the gait cycle). The left STN also demonstrated significant power decrease during right leg swing period, and beta band (13-30 Hz) decrease during right heel strike (Figure 1A and B, top).

**Figure 1.**
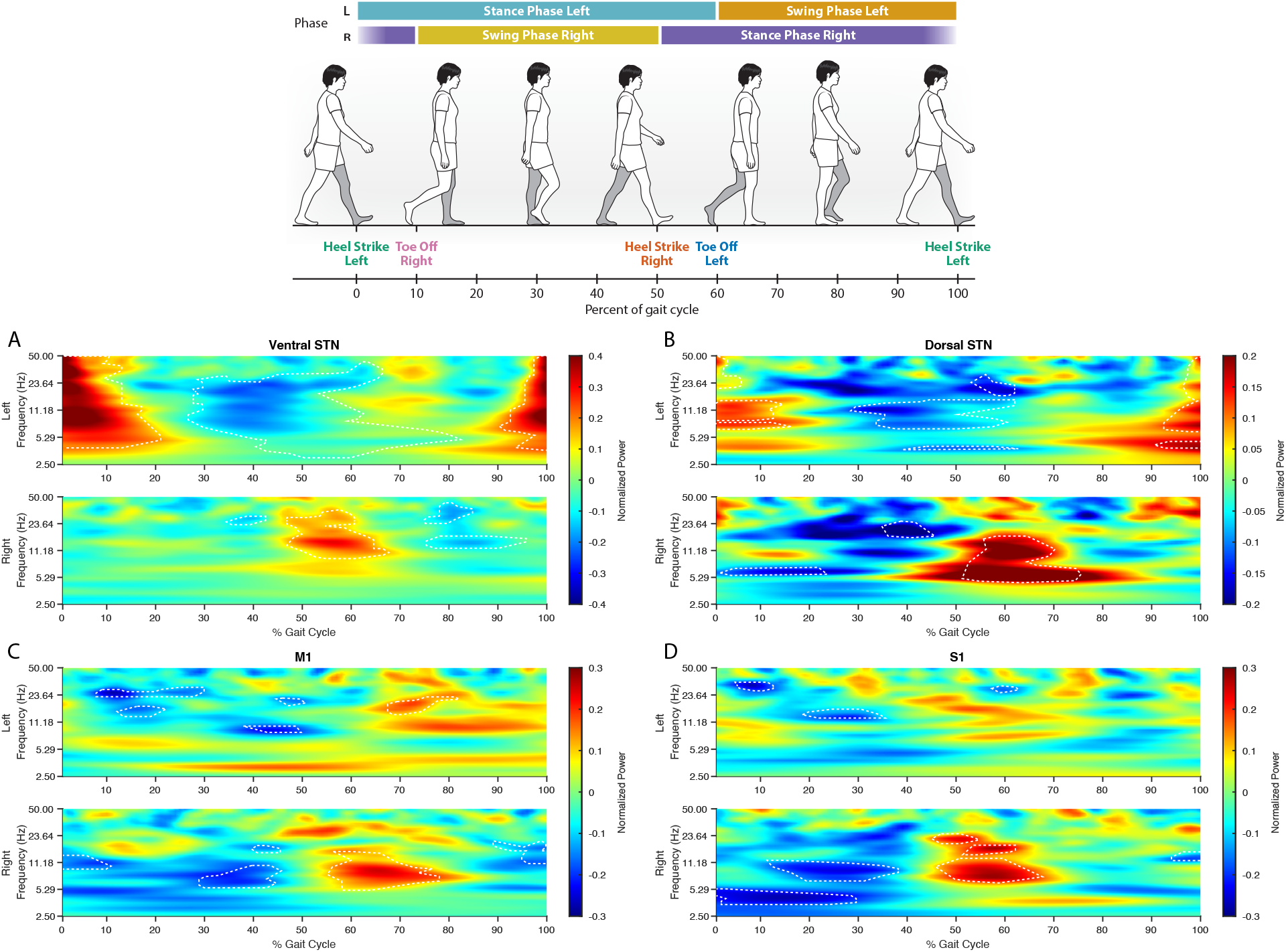
STN and cortical local field potentials show spectral power modulations during the gait cycle. Grand average z-score spectrograms from the dorsal and ventral STNs, M1, and S1 normalized to a gait cycle. (A and B) Significant power increases are seen during weight acceptance of the left leg in the left hemisphere (∼0-10% gait cycle) and right leg in the right hemisphere (∼50-60% gait cycle). Power increases was observed in a wide frequency band (10-50 Hz) in the ventral STN and in low frequency band (5-15 Hz) in the dorsal STN. Significant beta (13-30 Hz) desynchronization was also seen during contralateral leg swing and heel strikes. (C) Left M1 shows alpha (8-10 Hz) and beta desynchronization during right leg heels strike and initial right leg swing, respectively. Right M1 shows increased theta-alpha (5-12 Hz) during initial left leg swing and decreased beta around left heel strike. (D) Significant decreased beta power is seen during left leg weight acceptance and initial right leg swing. Increases in theta-beta power (5-23 Hz) were seen during weight acceptance of the right leg and initial left leg swing. (A, B, C, D) Gait cycle percentages and frequencies where power was significantly different compared to the average power during the entire walking task is outlined by the dashed white lines. A linear mixed-effect model was used to determine significance with p-value < 0.05.

M1 and S1 also demonstrated power fluctuations throughout the gait cycle, though the left and right hemispheres did not show coordinated, reciprocal changes. The left M1 showed decreased beta band amplitude during initial right leg swing (∼10-30% of gait cycle), decreased alpha band (8-10 Hz) power during right heel strike (∼40-50% of gait cycle), and increased beta band amplitude during right leg mid stance (∼65-80% of gait cycle) (Figure 1C top). The right M1 showed increased theta-alpha power during left leg initial swing and decreased beta band around left heel strike (Figure 1C, bottom). In S1, left and right hemispheres showed significant decreased power during weight acceptance of the left leg and initial right leg swing (Figure 1D). Additionally, the right S1 showed increased theta-beta (5-23 Hz) power during initial weight acceptance of the right leg and initial swing of the left leg (Figure 1D, bottom).

### STN interactions with motor and sensory cortices during different phases of the gait cycle

Because the STN has direct connections with sensorimotor cortices and plays important functions in motor control, we examined whether the STN interacts with the cortex during specific phases of the gait cycle. To determine the nature and degree of this interaction, we compared the averaged magnitude-squared coherence value between the ventral/dorsal STN and M1/S1 for each brain hemisphere during the gait cycle. We found increased coherence between STN and M1 during contralateral leg toe off and initial contralateral leg swing, similar to the power modulations seen in the ventral and dorsal STN (Figure 2). Dorsal STN-M1 and right ventral STN-M1 showed increased coherence in the theta band, while the left ventral STN-M1 showed increased coherence in the alpha/beta band. These increases in coherence were coordinated between the two hemispheres and were offset by half a gait cycle. The left STN-M1 coherence also demonstrated decreased beta coherence coinciding with right toe off (Figure 2A and B, top).

**Figure 2.**
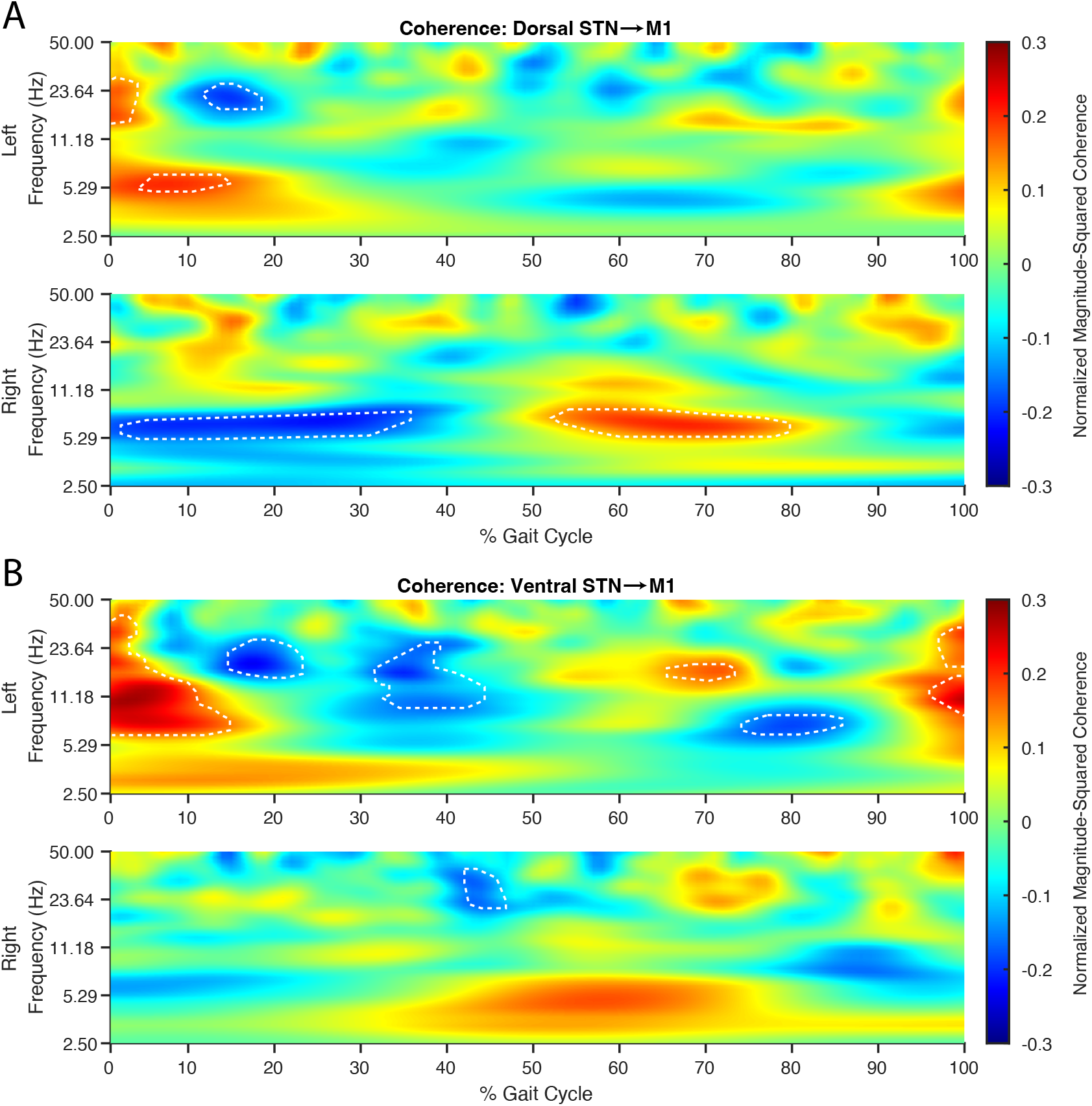
Low frequency STN-M1 coherence increase during the initiation of contralateral leg swing. Grand average z-score coherogram from dorsal and ventral STNS to M1 normalized to a gait cycle. Coherence modulation was seen in both hemispheres during weight acceptance of the ipsilateral leg and contralateral toe off. (A) Dorsal STN-M1 coherence showed significant increases in the theta band (5-8 Hz) during the initiation of contralateral leg swing through mid-swing. Additionally, the left hemisphere showed beta band coherence increases during initial ipsilateral weight acceptance and decreases during contralateral toe off. (B) Ventral STN-M1 coherence showed increased coherence was seen during contralateral toe off and contralateral leg swing. Left hemisphere showed these changes in the theta-beta band, while right hemisphere changes were in the theta band. (A and B) Gait cycle percentages and frequencies where coherence was significantly different from the average coherence during the entire walking task are outlined by the dashed white lines. A linear mixed-effect model was used to determine significance with p-value < 0.05.

For STN-S1 coherence, alternating increases in theta/alpha coherence was strongly seen between ventral STN to S1 coherence during the double support period, from ipsilateral heel strike to contralateral toe off (Supplementary Figure 1A). For dorsal STN to S1 coherence, this was not seen in the left STN-S1 (Supplementary Figure 1B, top).

We explored the phase lag during periods of high theta frequency coherence between M1 and STN across all gait cycles for all patients and did not find a consistent phase relationship between the two regions (Supplementary Figure 2). This suggests that the coherence modulations are driven primarily by cortical-subthalamic amplitude coherence during the gait cycle.

### Patient-specific oscillatory biomarkers of gait

Because our data showed several distinct gait-related frequency bands of modulation during the gait cycle, we used a data-driven approach to determine individual-specific frequency bands that are putative biomarkers for heel strike and toe off events. We created frequency bands of varying lengths ranging from 0-50 Hz, extracted power spectral density values at left and right heel strike and toe off events, and performed an ANOVA test for each band (Supplementary Figure 3). We found that each patient had unique frequency bands where power values significantly differentiated gait events (heel strike and toe off for each foot) (Figure 3). Significant gait-event-modulated frequency bands (*F3,varying* = 3.65-7.24; Table I) were found within all canonical frequency bands, with a majority in the theta and beta bands (Figure 3A, shaded areas). Frequency ranges of the gait-event-modulated bands varied depending on the patient and recording area but were typically a sub-range of the canonical bands; these frequency ranges are denoted in Figure 3A as the width of the shaded area. By comparing the instantaneous power spectral density during each of the four gait events, we found power differences between gait events that are temporally distinct in relation to the gait cycle (Figure 3A, inset plots). The left heel strike (green line) and right toe off (pink) events are temporally close to each other (within 10% of the gait cycle), similar to the right heel strike (orange) and left toe off (blue) gait events. Therefore, the gait events occurring in temporal proximity have a more similar power spectra profile compared to those of more temporally distinct gait events (i.e., left vs right heel strike) (Figure 3A).

**TABLE I.** ANOVA SUmmary

|  | Subject 1 | Subject 2 | Subject 3 |
| --- | --- | --- | --- |
| $DF_{bg}$ | 3 | 3 | 3 |
| $DF_{wg}$ | 195 | 383 | 688 |
| mean Significant F-Stat | 3.65 | 7.24 | 4.28 |

$DF_{bg}$ = Degree of Freedom Between Groups
$DF_{wg}$ = Degree of Freedom Within Groups
F-Stat = F-Statistic Value

**Figure 3.**
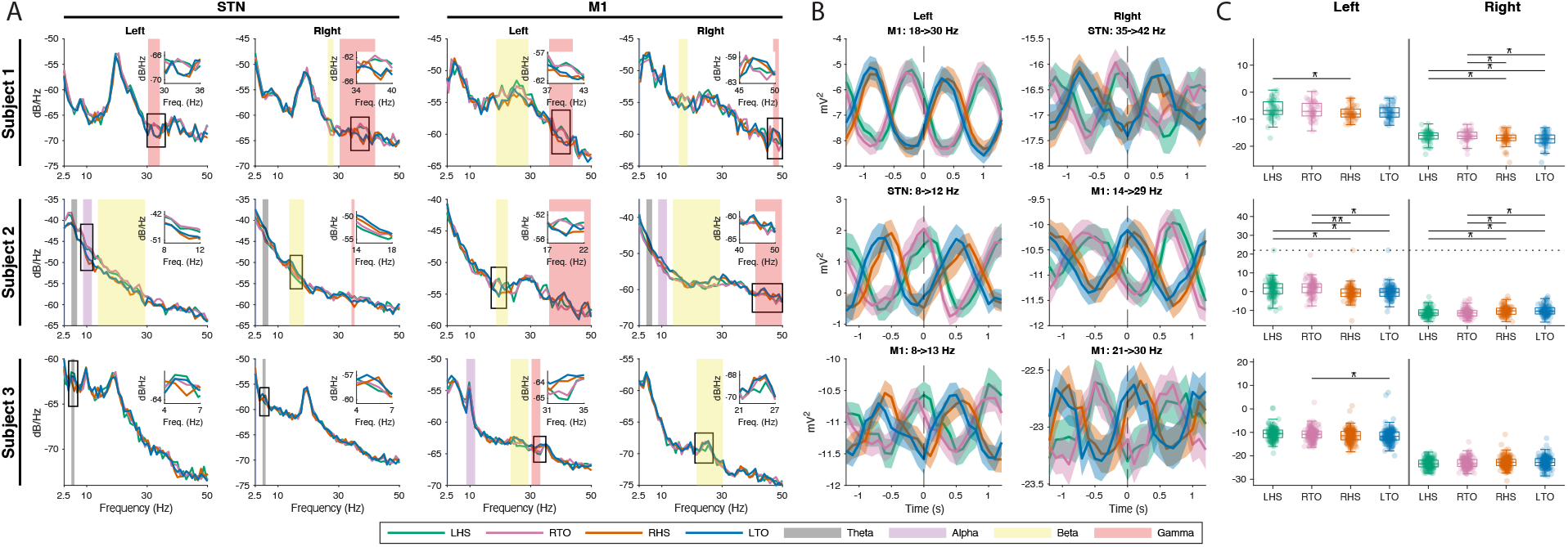
Unique frequency bands within each subject can differentiate gait events. (A) Average heel strike and toe off PSDs from the STN and M1. Each subject had unique frequency bands where power during heel strikes (left heel strike = green, right heel strike = orange) and toe off (left toe off = blue, right toe off = pink) gait events were significantly different (p < 0.05). The unique frequency bands were mainly found within the canonical frequency ranges (color of shaded area), but rarely spanned the entire range (width of shaded area). Inset plots show power differences between gait events temporally distinct from each other in relation to the gait cycle. (B) Average power and standard error ± 1 second around the gait event. Reciprocal out-of-phase power modulation is seen between temporally distinct gait events in all subjects. Furthermore, all left hemisphere data show higher power during left heel strike/right toe off and most of the right hemisphere data show higher power during right heel strike/left toe off. (C) Boxplot of gait event power within the frequency bands from B. Individual gait event powers are shown as transparent colored dots with outliers shown on the dotted line. Multiple comparison tests were performed against each pair of gait event within the same hemisphere. Level of significance is indicated as follows: * = p<0.05 and ** = p<0.005.

To evaluate how the amplitude of these individual gait-modulated frequencies change over time, we calculated the short time Fourier transform for each gait cycle and averaged the power for the gait-modulated frequency band ±1 second around each gait event. All averaged band power oscillated for the duration of the gait cycle (average gait cycle length ∼1 second) (Figure 3B). Power averages for the left heel strike (green) and right toe off (pink) events are antiphase to the right heel strike (orange) and left toe off (blue) events within each contact. In all subjects, the left and right hemispheres showed out-of-phase, reciprocal power modulations across different contacts. For instance, all subject’s left hemisphere cortical or basal ganglia LFPs showed higher power during left heel strike/right toe off and two subject’s right hemisphere contacts showed higher power during right heel strike/left toe off events.

To investigate whether each gait event’s instantaneous powers are distinct from each other, a multiple comparison test was performed between all possible pairs of gait events. Significant power differences were found between left and right heel strikes in subjects 1 and 2 in both hemispheres (Figure 3C, green vs orange). Other significant differences occurred between toe off events (Figure 3C, pink vs blue), and, occasionally, between heel strike and the contralateral toe off event. Gait events temporally close to each other did not differ in power.

### Decoding gait events based on cortical and subcortical LFPs

Given that our findings show that power differences exist between gait events, we investigated if we could decode gait events using these personalized “gait biomarkers.” Because our data has shown modulation during initial leg swing and toe off gait events characterizes switching from stance to swing phase, we built the classifiers to decode left and right toe off. We could classify toe off events with ≥68% accuracy in all subjects from at least one of the contacts, and that 61% of the trained models achieved significant above-chance accuracy (Figure 4). No one specific classification model outperformed the others consistently across subjects. Instead, the model that achieved the highest accuracy was subject specific. Additionally, some subjects gait event decoding accuracy was greater in models trained from the cortices contacts versus the STN. For subject 1, KNN had the highest accuracy (77-81%) for both brain hemispheres. For subject 2, the highest accuracy achieved by both brain hemispheres was 70%, but the model with this accuracy differed (left – XGBoost Tree; right – logistic regression). For subject 3, all models had similar accuracies across both brain hemisphere data. Overall, median model accuracies were greater than chance, i.e., 50%, and ranged from 56.4% to 61.8% (Table II). Further analysis of the models showed median discriminatory ability with area under the curve (AUC) values ranging from 0.583-0.663, while the best models achieved high discriminatory values (AUC > 0.84).

**TABLE II.** CLassification SUmmary

| Classification Model | Median Accuracy | Maximum Accuracy | Median AUC | Maximum AUC |
| --- | --- | --- | --- | --- |
| KNN | 0.564 | 0.808 | 0.606 | 0.882 |
| Logistic Regression | 0.583 | 0.765 | 0.583 | 0.847 |
| LDA | 0.577 | 0.706 | 0.587 | 0.847 |
| XGBoost Tree | 0.618 | 0.882 | 0.663 | 0.889 |

KNN = K-Nearest Neighbor, LDA = Linear Discriminant Analysis

**Figure 4.**
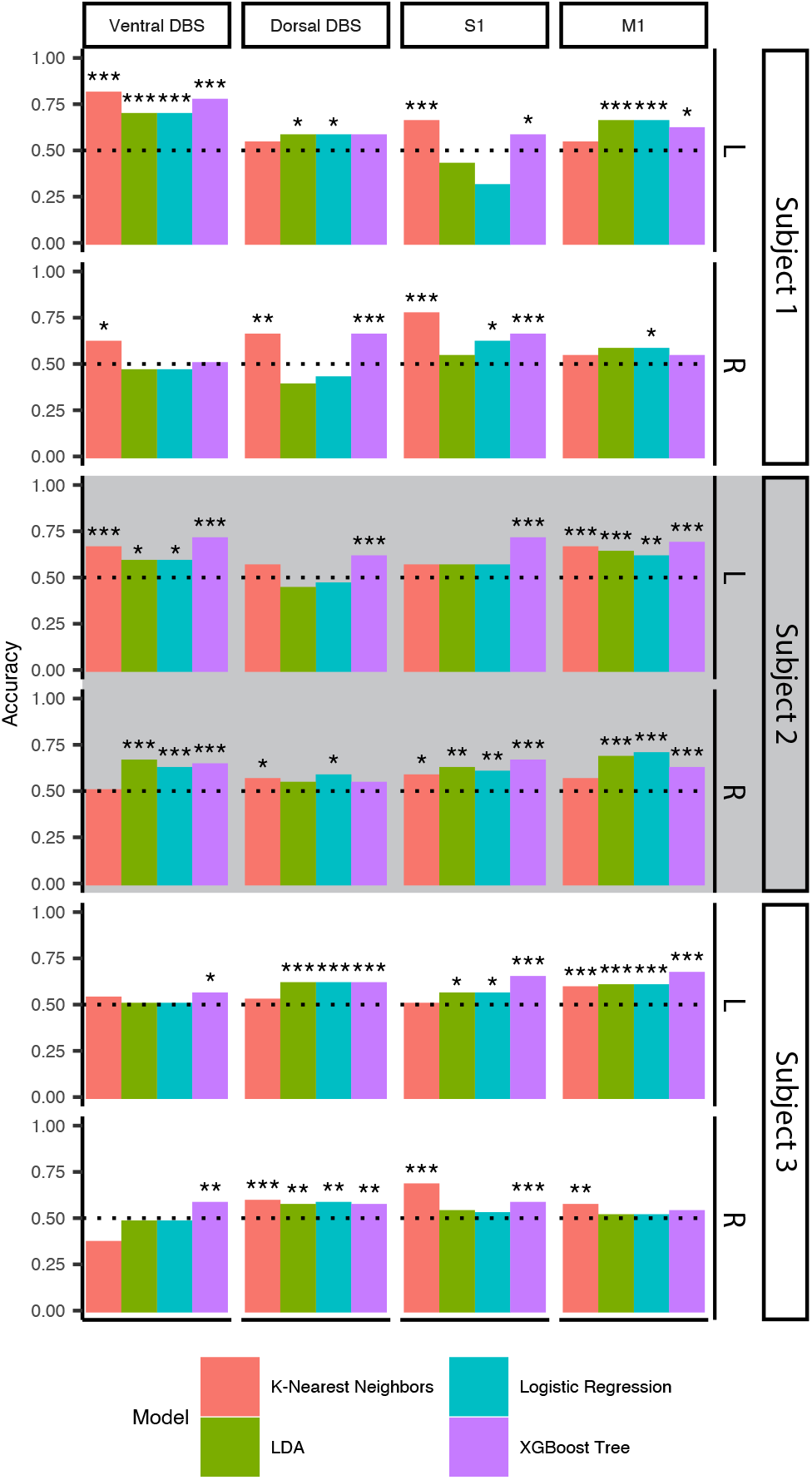
Gait event decoding using oscillatory features achieves greater than chance accuracy. Four ensemble classifiers were trained on left and right toe off events for each contact and hemisphere across all subjects. All subjects had at least one contact where at least one model’s classification accuracy was ≥68%. Maximum accuracy for all classification model types were between 70.6-88.2%. Maximum discriminatory ability was calculated using the area under the receiver operator characteristic curve and ranged between 0.847-0.889. Columns show recording area and rows indicate recording hemisphere and subject. Ensemble models were K-nearest neighbors (red), linear discriminant analysis (green), logistic regression (blue), and XGBoost Tree (purple). See also Table II.

Since STN-M1 coherence showed gait cycle-related changes, we also explored whether coherence between ventral and dorsal STN to S1/M1 could classify toe off events. Model training and significant testing mirror the process above with the exception that the data used to train the model were the coherence at left and right toe offs. Across all subjects and coherence pairs, we were able to decode left and right toe offs with ≥59% accuracy (Supplementary Figure 4). Between the STN and M1, the model with the highest accuracy was achieved by subject 1’s KNN model at 73%, which also achieved the highest AUC value of 0.71, indicating a medium discriminatory ability (Supplementary Figure 4, column 1 and 2). On average, models trained on coherence between dorsal STN and M1 had a higher accuracy (56.5% – dorsal versus 51.5% – ventral) and AUC values (0.58 – dorsal versus 0.52 – ventral) than models trained on coherence between ventral STN and M1. Between the STN and S1, the model that obtained the highest accuracy was subject 1’s XGBoost Tree model at 81% (Supplementary Figure 4, column 3 and 4). This model achieved an AUC value of 0.80, indicating good discriminatory ability. Overall, models built using coherence data between STN and S1 had higher accuracy, with a greater percentage of models able to significantly decode toe offs above-change (33% - STN/M1 versus 58% STN/S1).

## Discussion

We used chronic invasive recordings in PD patients to advance our understanding of dynamic subthalamic and sensorimotor oscillatory changes that underlie natural overground walking. First, we demonstrate the novel finding that STN displays increased low frequency (4-12 Hz) activity during the double support period prior to contralateral leg swing. Furthermore, STN shows increased theta frequency coherence with the primary motor during initiation of contralateral leg swing, implicating a potential mechanism for the supraspinal network to scale and fine tune leg muscle activation during stepping. Our findings support the hypothesis that oscillations from the basal ganglia and cortex direct out-of-phase, alternating power fluctuations between the two hemispheres in accordance with the gait cycle, which may indicate a mechanism to coordinate and maintain continuous bipedal locomotion in humans. In addition, we identified patient-specific, gait-related biomarkers in both subcortical and cortical areas at discrete frequency bands. Exploratory ensemble classification models showed above-chance accuracy in classifying left and right gait events using oscillatory power features. These putative biomarkers of the gait cycle may eventually be used as control signals to drive adaptive DBS to further improve gait dysfunction in patients with PD.

### Alternating multi-frequency modulations from bilateral STNs during gait

Several groups have described beta power modulations within the STN during the gait cycle between the left and right hemispheres during seated stepping (Fischer et al., 2018) and overground walking (Arnulfo et al., 2018; Canessa et al., 2020; Hell et al., 2018) in PD patients. Because elevated beta synchrony within the STN is associated with the akinetic state in PD, it is logical that beta desynchronization is required for movement, including gait. Beta activity has also been reported to have different power modulations between upper and lower extremity movement (Tinkhauser et al., 2019). We found that these gait-event related alternating power modulations between the left and right STNs are not limited to the high beta frequency range but also involve other low frequency bands.

What are the roles of subthalamic lower frequency (theta and alpha) modulation during gait? Previous studies on upper extremity movement tasks have shown event-related theta/alpha frequency synchronization within the STN at the onset and throughout the duration of movement, where sustained voluntary muscle contraction are required (Kato et al., 2016; Tan et al., 2013). In some cases, the amplitude of these theta/alpha oscillation correlate with the force generated during hand movement (Anzak et al., 2012). STN theta activity has been shown to have a role in the cognitive control of movement, such as during sensorimotor conflict (Aron et al., 2016; Zavala et al., 2017) and response inhibition (Alegre et al., 2013). We posit that these low frequency oscillations emerge from the STN during periods of gait that require greater cortical engagement. Based on increases in STN theta/alpha power we found during the transition from double support (both feet on the ground) to single support (ipsilateral leg on the ground) period, we postulate that these low frequency modulations engage multiple motor cortical areas to generate the appropriate scale and force required during contralateral leg swing to maintain stable single limb support and bipedal locomotion.

While some suggest that low-frequency modulations during gait may be secondary to movement-related artifacts (Hell et al., 2018), we believe that these low-frequency oscillations reflect physiological signals for several reasons. First, spectral activities that change during the gait cycle are focal in frequency range and are not broadband in nature (Figure 1). Second, the spectral power changes in left and right STNs are offset by half a gait cycle, unlike in the previous study where both STNs showed concurrent spectral power increases during the gait cycle regardless of laterality; the authors of this study discussed that their results may be movement related artifact (Hell et al., 2018). Finally, the dorsal and ventral STN, as well as M1 and S1 contacts show different time-frequency changes from each other during the gait cycle. Because the electrodes containing these four bipolar electrode pairs are connected to the same RC+S device, movement artifacts would affect all recording sites from the same brain hemisphere in a similar fashion.

One key question is whether these gait-related oscillatory modulations reflect physiological or pathological gait patterns. While our patients did not have overt gait abnormalities such as shuffling gait or freezing of gait, they performed the walking task on dopaminergic medication, which can affect oscillatory activity (Foffani et al., 2006; Ray et al., 2008). In a study involving patients with segmental dystonia without gait disorders, power modulations in the theta, alpha, and beta frequency around heel strike and toe off was reported from the GPi (Singh et al., 2011). Furthermore, the same study reported theta-alpha frequency band power modulation during early stance and swing phase of the contralateral leg, similar to our data. The conclusion from our results is that effective continuous gait is not associated with changes in a single frequency band in the basal ganglia. While we cannot rule out the presence of compensatory signals, we speculate that our results are an indicator of physiological gait, rather than pathological. The dynamic changes of oscillations across different frequency bands may provide a mechanism to coordinate and recruit different cortical and subcortical areas in response to changes in posture, balance, and forward momentum during walking.

### Cortical-subthalamic interactions during gait

As the cortex is more accessible by noninvasive methods, multiple EEG studies have been performed in healthy subjects and PD. In healthy subjects, the sensorimotor cortex has shown increased theta, alpha, and beta band power during the double limb support period and decreased alpha-beta power during leg swing which are offset by half a gait cycle between the two hemispheres (Bulea et al., 2015; Gwin et al., 2011). In PD, there may be exaggerated synchronization in the theta, alpha, beta, and gamma bands during walking compared to age-matched healthy-controls (Miron-Shahar et al., 2019), with increased theta over the vertex (Cz electrode) associated with freezing of gait (Shine et al., 2014). In a study involving simultaneous recording of STN LFPs and scalp EEG during walking in PD patients with freezing of gait, the authors found cortical-STN synchrony in 4-13 Hz during effective gait (Pozzi et al., 2019). It should be noted that patients in that study were navigating through a course and the greater synchrony may reflect a higher cognitive demand during walking due to the task.

The spatiotemporal specificity of field potentials captures by the permanently implanted cortical electrodes indicate distinct interactions between the STN and different cortical areas during gait. We demonstrated increased ventral STN-S1 coherence in the low frequency ranges (theta-alpha) during the double-support period between ipsilateral heel strike and contralateral toe off. We also found increased dorsal STN-M1 theta frequency coherence during contralateral toe off and early contralateral leg swing. These alternations in coherence are offset by half a gait cycle between the left and right hemispheres. To our knowledge, this is the first report of distinct patterns of STN-S1 and STN-M1 synchrony during human gait. We speculate that increased STN-S1 coherence during ipsilateral heel strike to contralateral toe off may represent sensory integration during the double support period as one prepares for leg swing. Increases in STN-M1 theta coherence then follows, during initiation of contralateral leg swing, which may allow the motor cortex to regulate the force of leg muscle activation required to drive forward stepping during gait. While these M1-STN interactions may represent normal recruitment of leg muscles during weight acceptance and transfer phase of the gait cycle, they may also represent compensatory mechanisms by which greater cortical activity is required to drive and maintain locomotion in PD.

### Gait event decoding and potential clinical significance

A key finding from our study was that for each patient, a unique range of frequencies were significantly differentially modulated corresponding to the various gait events. While these frequency bands often overlap canonical bands, they are usually narrower and span many different canonical frequencies. The variations among patients may be due to slight differences in electrode placement. While our results show greater than chance median accuracy and acceptable to excellent discriminatory ability, the models may be under-optimized for each subject. Attempts to optimize the models for each subject were restricted to performing 10-fold cross validation on hyperparameters of the random forest feature selection model, KNN, and XGBoost Tree model. Possible hyperparameter values to try, however, were restricted to a static set of values across all subjects. By constraining the set of possible hyperparameter values, possible values that would result in better accuracy and discriminatory ability for different subjects may have been missed. Additionally, the ratio of features (1770 total) compared to observations during feature selection can over-fit the model, leading to poor feature selection. Nonetheless, our study demonstrates the feasibility of distinguishing gait events based on cortical or STN LFP power.

One of the reasons to identify gait-specific biomarkers is to use them as control signals for closed-loop, also known as adaptive, deep brain stimulation (aDBS). While current continuous DBS (cDBS) can improve many motor symptoms of PD, it is often ineffective to alleviate gait disturbances given that it does not account for the rapid dynamic changes that occur with the gait cycle (Wang and Choi, 2020). The Summit RC+S system implanted in our subjects allow for aDBS in real time and utilizes LDA to detect different brain states using Fourier transform power within a frequency band (Ansó et al., 2022; Sellers et al., 2021). The aDBS feature of the Summit RC+S device has been successfully tested in single cervical dystonia patient (Johnson et al., 2021), and in PD patients (Gilron et al., 2021a, 2021b). Furthermore, the aDBS implemented differed between PD and the cervical dystonia patient, with slow changes in stimulation amplitude in PD patients (seconds to minutes) and rapid changes in the cervical dystonia patient (100s of milliseconds). Therefore, given that rapid stimulation changes have been successfully implemented and the results from our LDA gait event decoding, it is feasible to implement real time aDBS to rapidly change stimulation parameters to improve gait function in PD patients.

### Limitations

Our sample size is small due to invasive nature of these studies with investigational devices. Patients performed all tasks while on medication, which may affect beta power modulation. However, all STN LFPs during seated rest while on medication did show a spectral peak in the beta range. Due to variations in patient anatomy and electrode placement, M1 electrodes may capture field potentials from different regions of the precentral gyrus for each brain hemisphere. Each patient also had variabilities in their gait cycle (time spent in stance versus swing phase), therefore making comparison across patients challenging.

Another limitation of our study is that our results may be related to arm swing rather than leg movement. In a separate study involving a high-density subdural strip electrodes overlaying the hand knob area, we observed that the motor cortex is attuned to different limb movements in different frequency ranges (i.e., greater beta modulation during arm swing vs. greater theta modulation during leg movement; C. Starkweather, personal communications, 04/2022). Additionally, there is increasing evidence pointing to the existence of intermixed neural tuning of the whole body, including leg and foot movement, in the “hand knob” area of the precentral gyrus in humans (Willett et al., 2020; Zeharia et al., 2012). Future studies with additional cortical electrode coverage over the leg and arms can yield important information about STN’s interaction with other cortical regions (prefrontal, premotor, supplemental motor area) during gait.

### Conclusion

This study provides new insights on the role of subthalamic and sensorimotor oscillations play in human gait. Increased low frequency synchrony during the transition from double support to single leg support may provide a mechanism for the STN to integrate sensory information and engage the motor cortex to scale the appropriate motor response during stepping. Our data also support the notion that the STN and sensorimotor cortices contain patient-specific, gait-related frequency modulations that can be used to distinguish between left and right gait events. This knowledge has the potential to be integrated into adaptive neuromodulatory therapies to improve gait functions in patients with PD.

## Materials and Methods

### Subject and electrode placement

Three male subjects previously diagnosed with idiopathic PD with clinical indications for DBS surgery were enrolled at the University of California - San Francisco. Inclusion criteria included baseline off-medication Movement Disorder Society – Unified Parkinson’s Disease Rating Scale Part III (MDS-UPDRSIII) scores between 30 and 80, greater than 30% improvement in MDS-UPDRSIII on-medication, and absence of significant cognitive impairment (>20 Montreal Cognitive Assessment). Additional recruitment information, inclusion, and exclusion criteria are described in our prior work (Gilron et al., 2021a). All patients provided written informed consent to an IRB approved protocol (IRB 310759).

The mean age of the subjects was 53.7 ± 10.2 years and the mean disease duration was 6.7 ± 2.1 years (Table III). Subjects did not exhibit major gait impairments, as their MDS-UPDRS III postural instability and gait disturbance sub score on medication were between 1 (slight) to 2 (mild) (Table III). Gait kinematic measurements (heel strike and toe off) were captured using wearable sensors and were synchronized with the LFP data using continuous accelerometry data from both the DBS and external sensors, as well as stimulation pulses in the beginning and end of recordings.

**TABLE III.** SUbject DEmographics

| ID | Age Range/Sex | Disease Duration | DBS Target | UPDRS III Total Off-medication | UPDRS III Total On-medication | UPDRS III PIGD* On-medication |
| --- | --- | --- | --- | --- | --- | --- |
| Subject 1 | 40-48/M | 06 | STN | 41 | 14 | 2 |
| Subject 2 | 54-60/M | 09 | STN | 34 | 09 | 1<sup>ab</sup> |
| Subject 3 | 58-63/M | 05 | STN | 35 | 12 | 1<sup>a</sup> |

STN = Subthalamic Nucleus
\* Sum of items: 3.9, 3.10, 3.11, 3.12, and 3.13
a UPDRS III item 3.12 was not performed
b Severe right leg dystonia developed at higher peak dose

All subjects underwent bilateral placement of cylindrical quadripolar DBS leads into the STN (Medtronic model 3389, Medtronic, PLC), quadripolar cortical paddle into the subdural space over the motor cortices (Medtronic model 0913025, Medtronic, PLC) and investigational sensing implantable pulse generators (IPG) over the pectoralis muscles (Medtronic Summit RC+S model B35300R, Medtronic, PLC) as previously described (Figure 5) (Gilron et al., 2021a). The cortical electrodes in our study are placed over the “hand knob” area of the precentral gyrus (Figure 5A). Each RC+S device was connected to an STN DBS electrode and a cortical paddle from the same brain hemisphere.

**Figure 5.**
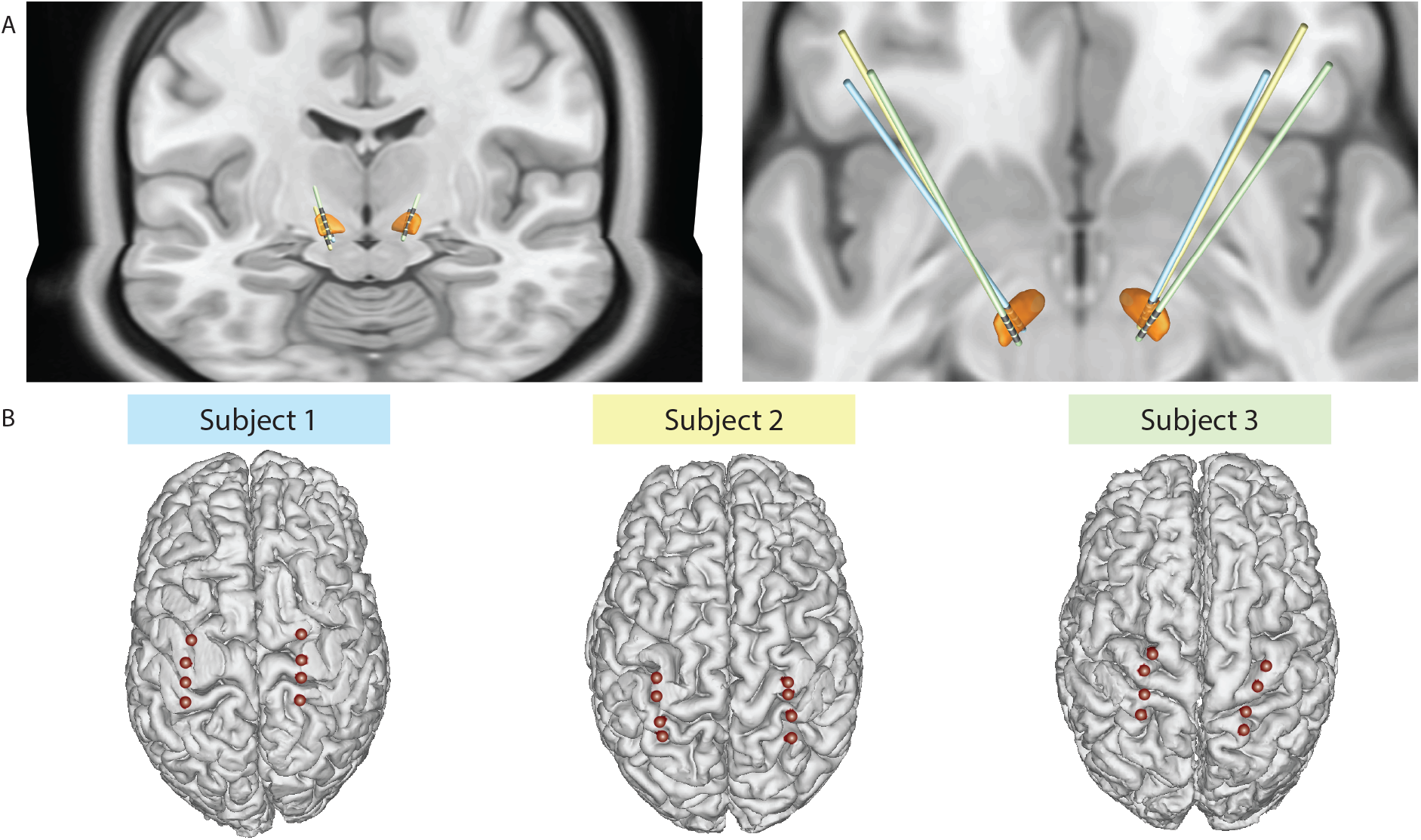
DBS and cortical lead localization. (A) 3D reconstructions of all DBS lead locations in the STN (orange). Individual subject’s leads are shown in by different colors. (B) 3D reconstructions of cortical electrode paddle location. The two most anterior contacts overlie the primary motor cortex (M1), while the two most posterior contacts overlie the somatosensory cortex (S1).

To anatomically localize STN DBS and cortical paddle contacts, postoperative CT images with electrode artifacts, were fused with preoperative T1-weighted MRI images. STN DBS lead reconstruction was performed semi-automatically in standard MNI space using the DISTAL atlas and TRAC/CORE algorithm available within Lead-DBS, an open-source MATLAB toolbox facilitating DBS imaging analyses (Ewert et al., 2018; Horn and Kühn, 2015). To reconstruct cortical paddle contacts we used the Intracranial EEG Anatomical Processing and Electrode Reconstruction Pipeline published in GitHub (to access code please visit: https://edden-gerber.github.io/ecog_recon/). Prior to fusing preoperative and postoperative images, T1 images were parcellated and converted into a standardized cortical surface mesh using FreeSurfer (Dale et al., 1999) and AFNI’s SUMA (Saad et al., 2004). Cortical contacts were then manually identified on the CT images in BioImage Suite (Papademetris et al.) and the electrode coordinates were projected onto the standardized mesh using a gradient descent algorithm in MATLAB.

### Gait kinematic measurements during natural walking

Two wireless sensor systems were used for gait phase detection and neural recording synchronization: Delsys Trigno® system (Delsys Inc. Natick, MA) and Xsens MVN Analyze (Xsens Technologies, The Netherlands). The Delsys sensors were from the Trigno and Avanti lines and included: two Avanti force sensitive resistor (FSR) adapters, two Avanti goniometer adapters, and two Trigno surface electromyography (EMG) sensors with a built-in accelerometer. The Avanti adapters were placed bilaterally on the shank of the leg, and the Trigno EMG sensors were placed over the skin on top of both IPG, and used for synchronization (see below). Attached to each FSR adapter were four pressure-sensitive footswitches (Delsys DC:F01), placed under the calcaneus, hallux, 1st metatarsal (1MT), and 5th metatarsal (5MT) on the sole of their shoes. A twin-axis digital goniometer (SG110/A) was placed next to the lateral malleolus and attached to a goniometer adapter. The Xsens system is comprised of 14 inertial measurement unit sensors placed over the entire body and limbs for wireless motion tracking. Subject 1 had both Delsys and Xsens systems attached, but FSR and goniometer data was not available. Subject 2 only used the Delsys system for collection of FSR and goniometer data. Subject 3 had both systems attached to determine gait events.

Once all sensors were placed, subjects walked overground in a straight path of at least 15 feet before turning around at their preferred walking speed for 2 minutes. Gait events illustrated in Figure 6B (left and right toe off and heel strike events) were determined using a custom MATLAB script. Heel strike (beginning of stance, end of swing) was defined as the time when the calcaneus or 5MT foot pressure crosses over a predefined threshold in the positive direction (Figure 6B dotted line). Toe off (end of stance, beginning of swing) was defined as the time when the hallux and 1MT foot pressure crosses over the threshold in the negative direction. A threshold of 5% of the maximum force detected was used. For subject 1, heel strike and toe off events were determined differently using ankle kinematic data from the Xsens system. Toe off was defined as the time of peak ankle plantarflexion velocity, while heel strikes was defined as the time of ankle velocity impulse. For all subjects, heel strike and toe off events were visually inspected to FSR or Xsens ankle accelerometry data, and erroneous events were manually corrected. Turns were excluded from analysis. 40 gait cycles were included for analysis from subject 1, 67 for subject 2, and 106 for subject 3. All subjects were on their typical dose of Parkinsonian medication during the task.

**Figure 6.**
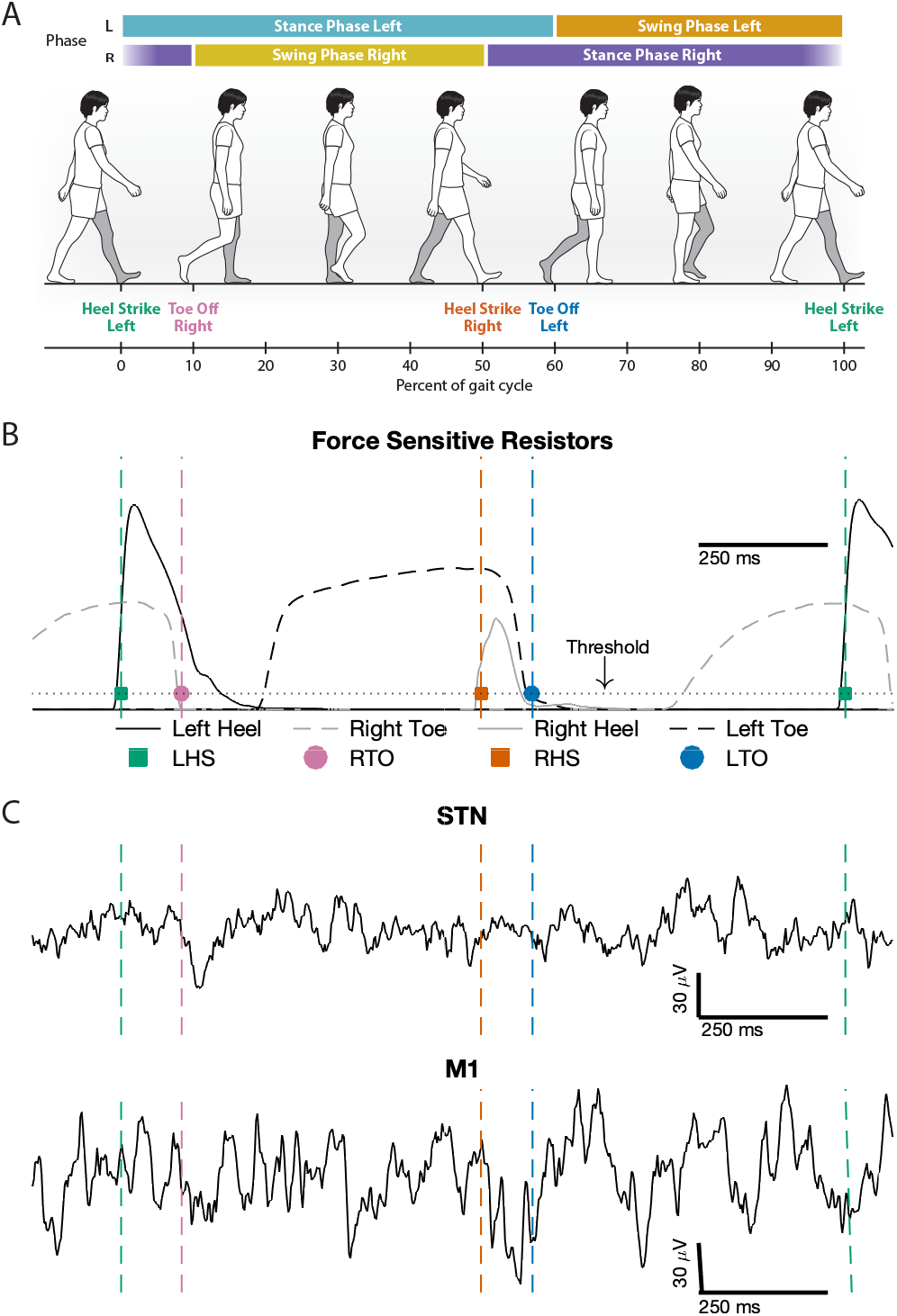
Synchronized gait kinematic data with raw local field potential recordings during natural walking. (A) Illustration of gait events and phases during a single gait cycle, aligned to left heel strike (0% gait cycle). (B) Heel strike (squares) and toe off (circles) gait events were detected from the left (black) and right (gray) force sensitive resistor data. Heel strikes were detected when the heel force (solid line) exceeded a threshold (dotted line), and toe offs were detected when toe force (dashed line) fell below the threshold. (C) Example local field potential recordings from both STN and M1 synchronized to a gait cycle.

### Neural recordings, data synchronization, and data processing

Local field potentials (LFPs) were streamed from two sets of STN and cortical electrodes pairs from each brain hemisphere. The two STN electrode pairs were recorded in the following configuration: +2-0 and +3-1, where contact 0 is in the ventral STN, contact 3 just above the dorsal border, and contacts 1 and 2 in the motor territory based on microelectrode recording identifying movement-related cells. +2-0 will be referred to as ventral STN and +3-1 as dorsal STN (Figure 5A). The two cortical electrode recording configuration were +9-8 (primary somatosensory cortex; S1) and +11-10 (primary motor cortex; M1), based on reversal of somatosensory evoked potential (Swann et al., 2018) and imaging reconstruction. LFPs were sampled at 500 Hz. Hardware pre-amplifier high-pass and low-pass filters were set to 0.85 Hz and 450 Hz, respectively, with an additional hardware low-pass filter at 1700 Hz after amplification. Accelerometry data from the Summit RC+S system was also collected and sampled at 64 Hz from all subjects. The Summit RC+S system recording capabilities have been previously described (Gilron et al., 2021a).

LFPs and accelerometry data were transmitted wirelessly from the RC+S device to a data collection computer in JavaScript Object Notation (JSON) format and saved for offline analysis. Once the recordings finished, the JSON data were analyzed to extract relevant timing and LFP data using open-source code (https://github.com/openmind-consortium/Analysis-rcs-data) built specifically to interface with the Summit RC+S JSON data.

The extracted neural and accelerometry data from the RC+S device were thereafter synchronized with all external sensor data via two methods: 1) alignment based on acceleration, by aligning acceleration peaks captured by the RC+S, Delsys Trigno sensor, and the Xsens accelerometer sensors, and 2) alignment of the brief stimulation pulses at the beginning of the task (10 seconds of brief burst of 5 Hz monopolar stimulation with the IPG case as the anode and contact 1 on the DBS lead as the cathode delivered at 2 mA) captured by LFP and surface EMG data.

### Spectral Analysis

Four signal processing methods were applied to each neural recording using built-in MATLAB functions: continuous wavelet transform (CWT), wavelet coherence, short-time Fourier transform (STFT), and power spectral density (PSD). We used wavelet transformation because it has greater low-frequency resolution, which is of interests given that our data showed low frequency power modulations during gait. We also used the Fourier transform because this is the spectral decomposition method used by the Summit RC+S system (Sellers et al., 2021).

The CWT was calculated using the function “cwt,” and wavelet coherence was calculated using the “wcoherence” function. The “spectrogram” function was used to calculate the STFT and PSD (1 second window length, 90% overlap, and transform length of 512 data points). These settings resulted in a STFT and PSD frequency resolution of 0.9766 Hz and a time resolution of 100 ms.

The CWT and wavelet coherence were used to calculate the grand average gait cycle spectrogram and magnitude-square coherogram from all recorded areas and all gait cycles across subjects. Spectrogram data for each LFP channel during individual gait cycles (when two same side heel strikes were within 2 seconds of each other) were extracted and divided into 100 gait cycle time bins. Power and magnitude-squared values were averaged across all gait cycles and for each subject and bin. Average values are z-score normalized (mean/standard deviation) to the average value during the entire walking period for each frequency and in each subject. Normalized values were averaged across subjects for each frequency and bin.

STFT was used to identify frequency bands where power differed between gait events. Power values at each gait event (left and right toe off and heel strike) were extracted from each brain hemisphere and contact pair. Then, all possible frequency bands were created between 0-50 Hz; a total of 1770 frequency bands were created. For each one of these frequency bands, the instantaneous power for each gait events were extracted. A one-way ANOVA test was used to compare the four different gait events within a frequency band for each recording location; the gait event was used as the grouping variable. Finally, a multiple comparison test was performed to determine which pairs of gait events had a significant difference within each frequency band. P-values were adjusted using Tukey’s Honest Significant Difference method.

Frequency bands with p-values < 0.05 were designated as gait-event-modulated frequency bands. To demonstrate power modulation within these gait-event-modulated frequency bands throughout the gait cycle, we extract the power around ±1 second of each gait event was extracted, and the mean and standard error were calculated.

### Gait event classification

Four classification model were built for each subject, brain hemisphere, and recording area to predict gait events from neural data and the coherence between the STN and M1. To enhance the stability and accuracy of the predictor models, ensemble learning with multiple machine learning algorithms was used (Polikar, 2006; Wolpert, 1992). The four ensembles consisted of a Random Forest (RF) model for feature selection model and one of the following classification models: K-nearest neighbors (KNN), logistic regression, linear discriminant analysis (LDA), and XGBoost Decision Trees (XGBoost). RF was used for feature selection as it has been shown to achieve better performance than other feature selection methods (Chen et al., 2020), and is robust to collinearity (Genuer et al., 2010). The four classification models were chosen to explore a range of different types of models and their assumption. All models were built in R with the “Tidymodel” framework (Kuhn and Wickham, 2020).

Features in the RF model were either instantaneous power or magnitude-squared coherence during toe off events in all possible frequency bands between 2.5-50 Hz, as we did not see significant power modulation above 50 Hz with the gait cycle and to remove low frequency bands that may be influence by the natural gait cycle frequency (i.e., ∼1-2 second per gait cycle). Power values were taken from the STFT, while magnitude-squared coherence values were taken from the output of the “wcoherence” MATLAB function. These frequency bands were extracted from all neural contacts which resulted in 4900 power features and 990 coherence features. All features were normalized to a mean of 0 and a standard deviation of 1. Prior to feature selection, hyperparameters, including the number of decision trees and number of features a tree considers during node splitting, of the RF model were optimized using 10-fold cross-validation with each data set stratified by toe off classes such that approximately equal numbers of left and right toe offs were in each data set. Once optimized, the RF feature selection model was trained on all normalized features. The “ranger” (Wright and Ziegler, 2017) package in RStudio (www.rstudio.com) was used for this analysis.

The top ten features with the largest variable importance value based on “permutation importance” (Altmann et al., 2010) were used to generate new data sets for each subject and brain hemisphere. Next, the new data sets were stratified and split with 75% of the data used for training and 25% of the data used for testing the classification models. The training data was further split into 10 folds for cross-validation hyperparameter optimization for the KNN and XGBoost models. Specifically, the number of neighbors to consider for classification was optimized for KNN. For XGBoost, two hyperparameters were optimized, the number of features that are randomly selected for each split in a decision tree and the minimum number of data points a node in a decision tree must have before increasing the tree depth. Following optimization and training, the accuracy (proportion of correct prediction) and receiver operator characteristic area under the curve (AUC) were calculated for each model using the test data set.

### Statistical Analysis

We examined differences between the power or magnitude-squared coherence at different frequencies during the gait cycle to the averaged values across the entire gait cycle at the same frequency. Data were included in a linear repeated-measure mixed model because this modeling approach has been shown to handle unbalanced number of observations (e.g., different number of gait cycles for each subject). A single fixed effect was use in the model, “source,” which represented whether the value was from a certain gait cycle percentage versus average across the entire gait cycle. Individual subjects were added to the model as a random to account for individual baseline neural power differences. Significance was tested using F-tests with Satterthwaite’s degrees of freedom method.

Statistical analysis of toe off decoding models were performed only on models that achieved greater than chance accuracy, i.e., ≥ 50%, given two classes. Significance was tested by permuting the toe off class labels and calculating the class accuracy on the permuted data. This procedure was performed 1000 times, and the number of permuted data sets where decoding classification accuracy reached or was greater than the non-permuted data set were counted. Models were determined to be significant if the counted number was < 5% of total number of permutations, i.e., < 50 permuted data sets (Herrojo Ruiz et al., 2014).

## Data Availability

All data produced in the present study are available upon reasonable request to the authors.

## Data availability

Data and analysis code are available upon reasonable request. Inquires can be sent to the corresponding author.

## Acknowledgements

The authors would like to thank our patients for participating in this study.

