## Supplemental Figures for "Decoding natural gait cycle in Parkinson’s disease from cortico-subthalamic field potentials"

### Supplementary Information

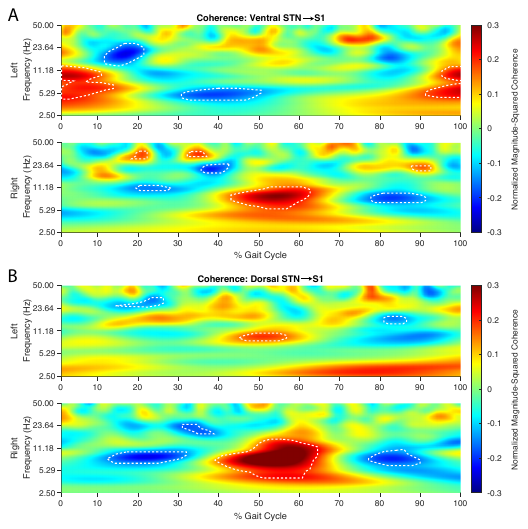

**Supplementary Figure 1 STN-S1 coherence show theta-alpha frequency increase during double support period between ipsilateral heel strike and contralateral toe off.** (A) Ventral STN-S1 coherence modulation was seen theta/alpha band across both hemispheres during double support period, the time between ipsilateral heel strike to contralateral toe off. (B) Increased theta/alpha coherence between dorsal STN to S1 was seen during double support period in the right hemisphere only. Left hemisphere also showed increased theta/alpha coherence during contralateral heel strike. (A and B) Gait cycle percentages and frequencies where coherence was significantly different from the average coherence during the entire walking task are outlined by the dashed white lines. A linear mixed-effect model was used to determine significance with p-value < 0.05.

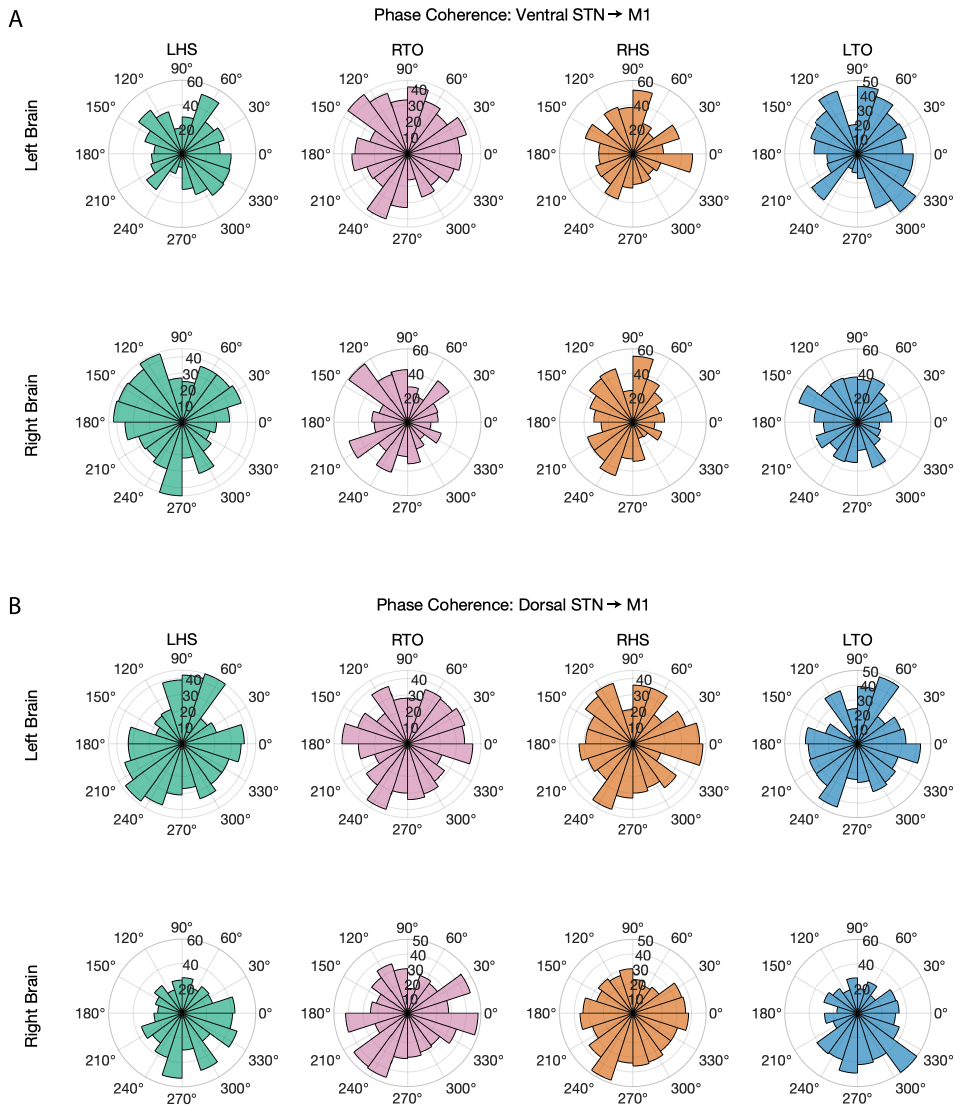

**Supplementary Figure 2 STN-M1 does not show consistent low frequency phase coherence during different gait events.** Polar histograms of the low frequency (5-6 Hz) coherence phase at different gait events across all subjects. (A and B) Phases at all gait events and brain hemispheres were not consistent and varied across the entire range of phases. LHS = left heel strike; RTO = right toe off; RHS = right heel strike; LTO = left toe off.

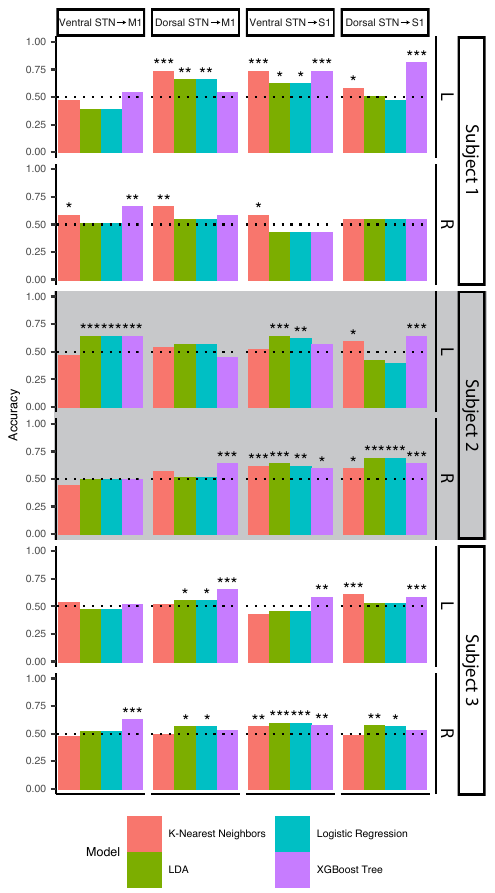

**Supplementary Figure 3 Example arbitrary frequency bands and ANOVA testing.** Related to Figure 5. Varying length frequency bands were created between 0-50 Hz. Each frequency is referenced as a bin. Start and end bin refers to the varying length frequency band’s start and end frequency. Power during left and right heel strike and toe off events were extracted from each frequency band and an ANOVA test was performed. The p-value of the ANOVA test was stored and a heat map was created. Example of the resulting heat map is shown from subject 2 M1 recorded area. Significant ANOVA test outcomes can be observed to fall within the low gamma band (30-40 Hz) frequency.

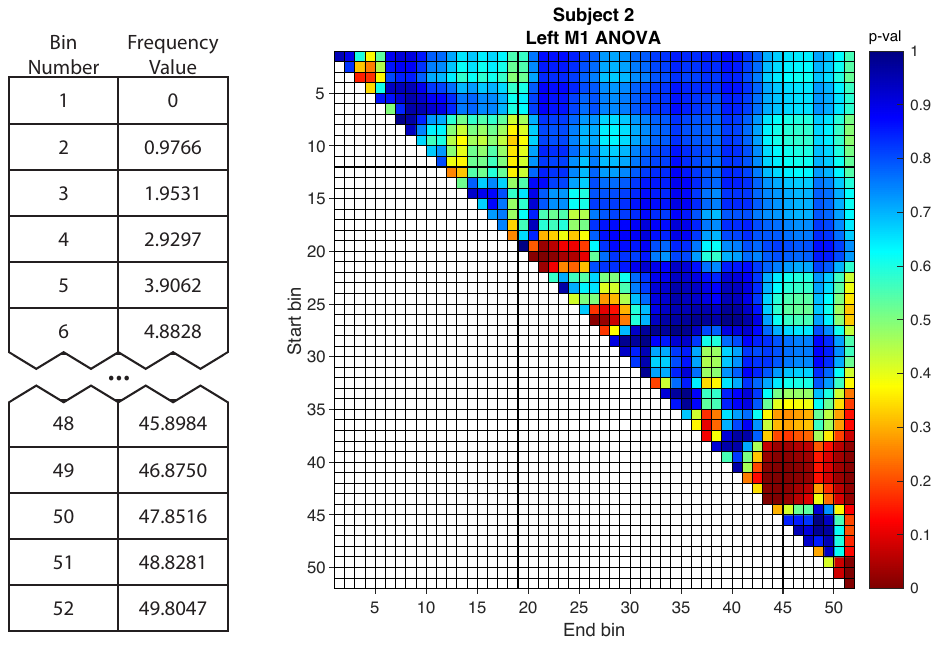

**Supplementary Figure 4 Toe off gait event decoding using STN-M1 coherence.** Four ensemble classifiers were trained using coherence magnitude squared values between the ventral and dorsal STN to M1 and S1. All subjects had at least one contact where toe offs were decoded with at least 59% accuracy. Overall, models trained on coherence between the STN and S1 had higher accuracy, and a greater number of models able to significantly decode toe offs compared to models trained on coherence between STN and M1.
